# COVID-19 mortality in Italy varies by patient age, sex and pandemic wave

**DOI:** 10.1101/2021.10.01.21264359

**Authors:** Francesca Minnai, Gianluca De Bellis, Tommaso A. Dragani, Francesca Colombo

## Abstract

**Background:** SARS-CoV-2 has caused a worldwide epidemic of enormous proportions, which resulted in different mortality rates in different countries for unknown reasons.

**Aim:** We aimed to evaluate which independent parameters are associated with risk of mortality from COVID-19 in a series that includes all Italian cases, ie, more than 4 million individuals infected with the SARS-CoV-2 coronavirus.

**Methods:** We analyzed factors associated with mortality using data from the Italian national database of SARS-CoV-2-positive cases, including more than 4 million cases, >415 thousand hospitalized for coronavirus disease-19 (COVID-19) and >127 thousand deceased. For patients for whom age, sex and date of infection detection were available, we determined the impact of these variables on mortality 30 days after the date of diagnosis or hospitalization.

**Results:** Multivariable Cox analysis showed that each of the analyzed variables independently affected COVID-19 mortality. Specifically, in the overall series, age was the main risk factor for mortality, with HR >100 in the age groups older than 65 years compared with a reference group of 15-44 years. Male sex presented an excess risk of death (HR = 2.1; 95% CI, 2.0–2.1). Patients infected in the first pandemic wave (before 30 June 2020) had a greater risk of death than those infected later (HR = 2.7; 95% CI, 2.7–2.8).

**Conclusions:** In a series of all confirmed SARS-CoV-2-infected cases in an entire European nation, elderly age was by far the most significant risk factor for COVID-19 mortality, confirming that protecting the elderly should be a priority in pandemic management. Male sex and being infected during the first wave were additional risk factors associated with COVID-19 mortality.

## Introduction

COVID-19, the disease caused by severe acute respiratory syndrome coronavirus 2 (SARS-CoV-2), is responsible for a worldwide pandemic of enormous proportions, in terms of the numbers of both infections and deaths. SARS-CoV-2 infection may be asymptomatic or symptomatic, with severity ranging from a few flu-like symptoms to severe respiratory manifestations requiring supplemental oxygen and to multi-organ failure [1–3]. COVID-19 mortality has been widespread throughout the world, but with considerable temporal and geographical variations [4–7]. Although the reasons for these differences are not yet fully elucidated, according to a meta-analysis[8] the predictors of mortality of COVID-19 that have been repeatedly observed in different case series from different countries are advanced age, male sex, and pre-existing comorbidities; in addition, some abnormal values of laboratory biomarkers have been associated with poor prognosis.

Italy was the first country after China to be heavily affected by the pandemic, and it has registered excess overall mortality and more than 127 thousand COVID-19-related deaths by July 25, 2021 [9,10]. The trends of cases and deaths, in most countries, have not been constant over time, but have fluctuated, with peaks of incidence called “waves”. The first wave, in Italy and other European countries, began in January 2020 and lasted until summer 2020 [7,11]. Indeed, during the summer there were few cases and deaths, but since the fall the numbers of cases and deaths rose again with subsequent waves [9], due in part to virus variants.

There is ongoing discussion about the effects of different waves on mortality in COVID-19 patients [11]. In particular, a few studies showed that mortality decreased after the first wave (i.e. after June 2020) [12,13], but these results have not yet been confirmed by the analysis of country-wide mortality data. Therefore, we analyzed factors associated with COVID-19 mortality in the Italian national case series.

## Materials and Methods

Data on people infected with SARS-CoV-2 in Italy (Italian COVID-19 epidemiological surveillance data) were obtained from the Italian Istituto Superiore di Sanità (ISS) after filling a request on April 15, 2021, at https://www.iss.it/richiesta-dati-covid19. The series included people whose PCR diagnosis of infection or first symptoms were recorded from January 28, 2020, to July 25, 2021. The data regarded each person’s age at diagnosis, sex, nationality, date of PCR-confirmed diagnosis, presence/absence of symptoms, date of hospitalization (if pertinent), the alive/dead status on the day of data sharing (i.e., July 25, 2021) and, if relevant, the date of death. Data on comorbidities were not provided.

### Survival analyses

Survival was analyzed in the entire cohort and, separately, for non-hospitalized and hospitalized patients. For the entire cohort and for non-hospitalized persons, we assessed mortality within 30 days of the date of infection detection (positive PCR test). Instead, for hospitalized patients, we assessed mortality within 30 days of the date of hospitalization. In all analyses, we considered only those persons for whom complete data were available regarding age, sex, date of infection detection or of hospitalization (depending on the group), and status (alive or dead) after 30 days. Patients who died on the day of infection detection or hospitalization were assigned 1 day of follow-up. Patients whose date of death was erroneously reported in the dataset as being before the date of infection detection or hospital admission were excluded from analysis.

Survival analyses considered the effects of age, sex and pandemic wave. In the choice of age groups, we took as reference the age group 15-44 years, instead of the youngest class (0-14 years). The reasons for this decision were that the age group 15-44 years has undergone only marginal alterations in overall mortality during the pandemic in European countries [14], and it is larger than the younger age class (0-14 years). Therefore, its use as reference provides good stability for risk estimates. For pandemic waves, we defined the first wave as that from the beginning of the dataset (January 28, 2020) until June 30, 2020, and the second wave as that from July 1, 2020, to July 25, 2021.

Associations between demographic-clinical features (i.e. age, sex and pandemic wave) and survival were evaluated in a Cox proportional hazard model using the survival package in R environment. The same package was used to draw Kaplan-Meier curves and run the log-rank test. The variables that were found to impact upon survival in univariable Cox analyses (with *P* <0.05) were analyzed in a multivariable Cox analysis. Cox and log-rank test *P* values <0.05 (two-sided) indicated statistical significance.

## Results

The Italian cohort of SARS-CoV-2-positive individuals included 4,333,014 persons of median age 46 years, with a slight predominance of female cases (**Table 1**). Overall, 50.4% of the cases were symptomatic and 415,390 were hospitalized, with 13.7% of them requiring intensive care. The median age of not-hospital patients was much lower than that of hospitalized cases (44 vs. 70 years). For the entire case series, 240,850 were diagnosed during the first wave, while 4,012,584 became infected after June 30, 2021. At the end of the study, 127,524 subjects were dead, with 35,837 having been diagnosed as infected before June 30, whereas 89,875 dead patients had been diagnosed after this date (not shown; for 1,812 subjects, information about the date of infection detection was missing). Over 31 thousand not-hospitalized people had died by July 25, 2021, while the total number of deaths among the hospitalized people was 95,907, of which 25,320 during the first wave. The median age at death of not-hospitalized patients was 86 years, with 94% of them being ≥65 years old (not shown). Instead, the median age of deceased hospitalized patients was 81 years, with 90% of them ≥65 years old (not shown). Surprisingly, more than 38 thousand people who were hospitalized were listed in the dataset as being asymptomatic.

**Table 1.**
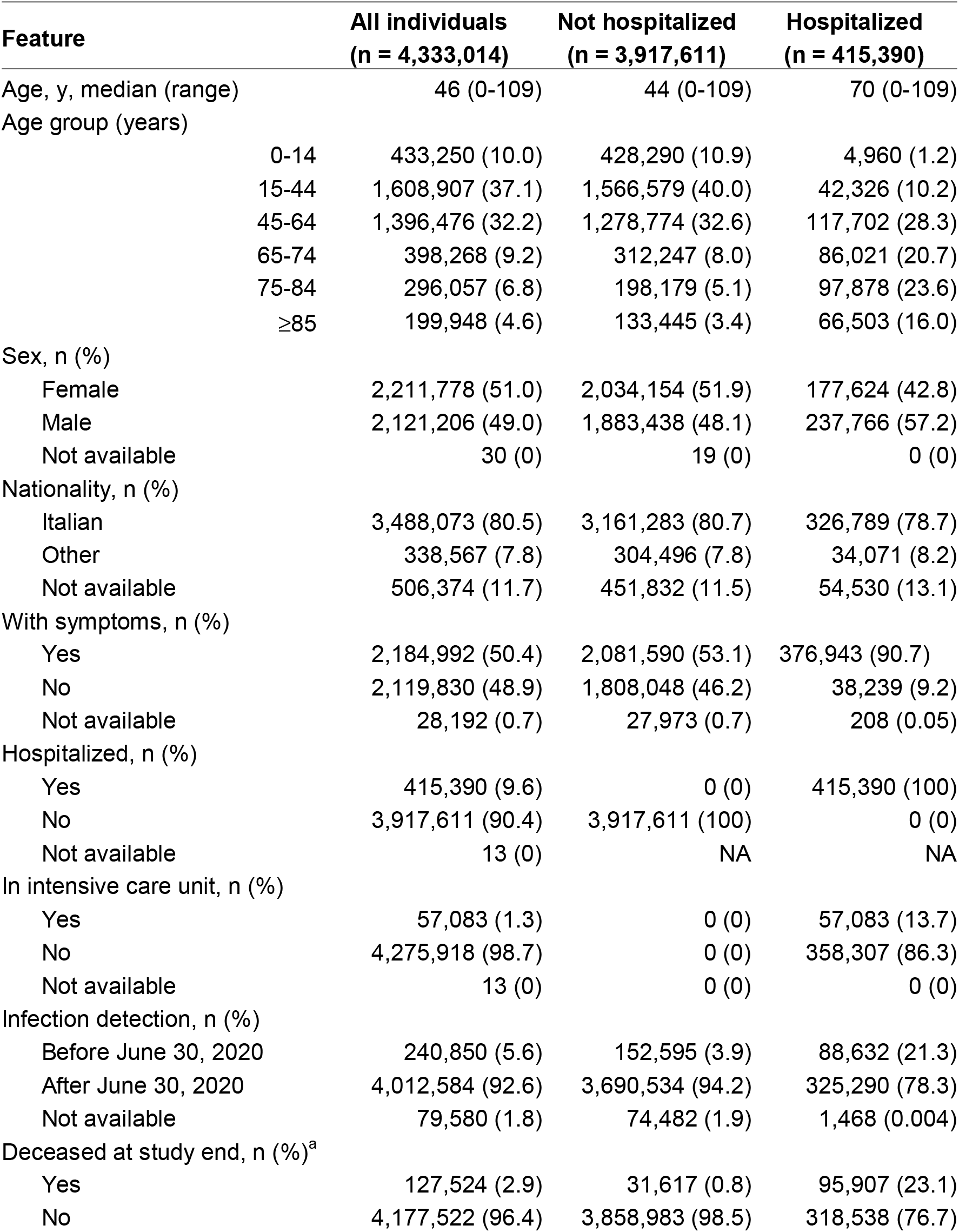
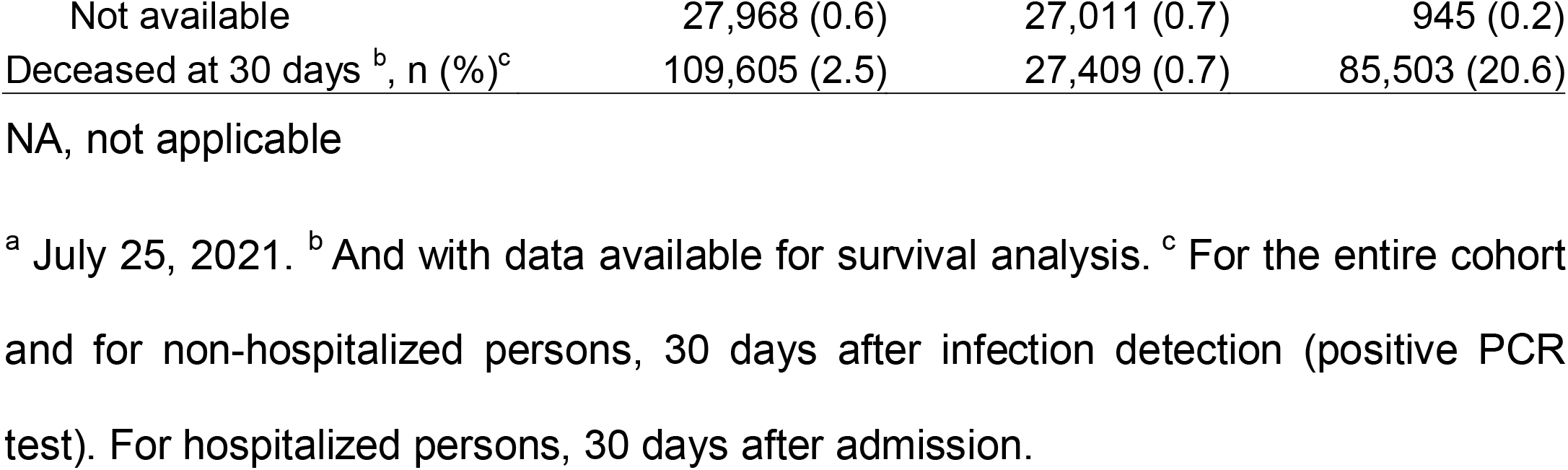
Demographic and clinical features of 4,333,014 SARS-CoV-2-positive individuals in the Italian national database, from January 28, 2020 to July 25, 2021.

To investigate the factors affecting mortality after SARS-CoV-2 infection, we first carried out univariable Cox analyses in the whole series and, separately, for non-hospitalized and hospitalized patients. For the whole series, the analysis was limited to 4,224,698 persons, after eliminating 108,316 persons with incomplete data (including 1,744 persons for whom the date of death was erroneously listed as being before the date of diagnosis or hospitalization). For non-hospitalized and hospitalized persons, these numbers were 3,816,311 and 412,942, respectively. We tested the effects of age, sex, and pandemic wave (first wave, before June 30, 2020, vs. later) on the risk of death 30 days after infection detection or hospitalization. Statistically significant associations (log-rank test, *P* < 2 × 10^-16^) for all three variables were observed, in the whole series and in the two subsets (**Figure 1**). The probability of survival decreased with increasing age and was lower for males than females and for persons who were diagnosed in the first wave than later.

**Figure 1.**
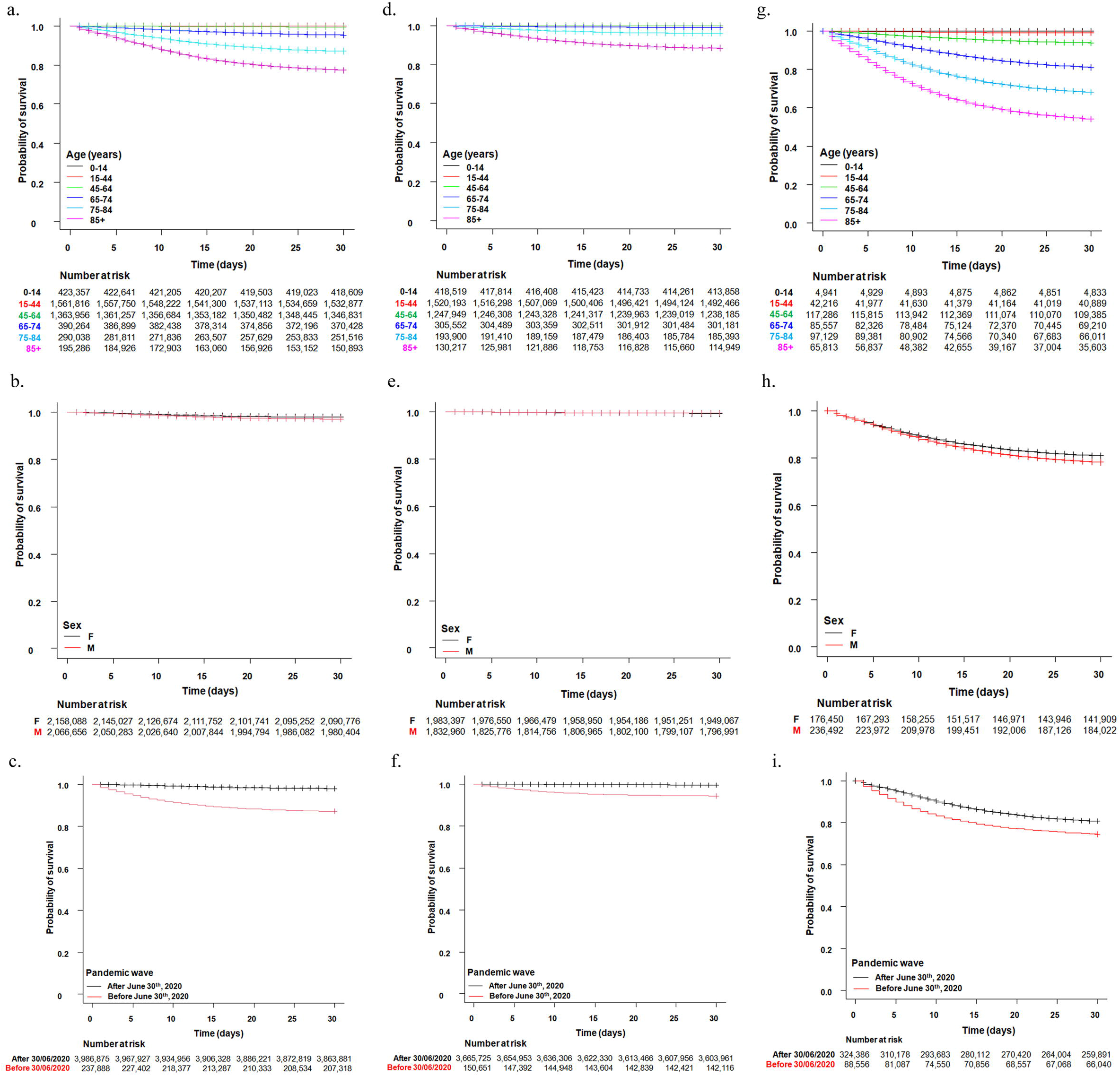
Kaplan–Meier survival curves for the entire Italian series of SARS-CoV-2-positive persons (panels A-C), for non-hospitalized persons (panels D-F), and for hospitalized COVID-19 patients in the Italian national COVID-19 series, by age group, sex, and pandemic wave (before vs. after June 30, 2020). Crosses denote censored samples. Numbers of patients at risk for each group, at each time point, are reported under each plot. Log-rank test *P* < 2 × 10^-16^, except for panel E for which *P* = 1.2 × 10^-11^.

Multivariable analysis of mortality in the whole series demonstrated that age, male sex, and the first pandemic wave were independent risk factors for death (**Table 2**). This analysis showed a sharp increase in mortality risk with increasing age, especially in persons ≥65 years old and with a tremendously high risk (hazard ratio [HR] = 692.4; 95% CI, 636.2 to 753.6) in the age ≥85 years group. Children (0-14 years old) showed a much lower risk of death (HR = 0.1) than both the reference age group and the older groups. Male sex was associated with a ∼2-fold risk of death (HR = 2.1), compared to females. The first pandemic wave (before June 30, 2020) was associated with an excess risk of death of almost 3-fold (HR = 2.7), compared to the subsequent period.

**Table 2.** Factors associated with death in the whole Italian series of people infected by SARS-CoV-2^1^

| Feature | Hazard ratio (95% confidence interval) | P-value <sup>2</sup> |
| --- | --- | --- |
| Age group (years) |  |  |
| 0-14 | 0.1 (0.1 – 0.2) | <2 x 10<sup>-16</sup> |
| 15-44 | 1 |  |
| 45-64 | 17.0 (15.6 – 18.5) | <2 x 10<sup>-16</sup> |
| 65-74 | 126.4 (116.1 – 137.7) | <2 x 10<sup>-16</sup> |
| 75-84 | 357.5 (328.5 – 389.1) | <2 x 10<sup>-16</sup> |
| ≥85 | 692.4 (636.2 – 753.6) | <2 x 10<sup>-16</sup> |
| Sex |  |  |
| Female | 1 |  |
| Male | 2.1 (2.0 – 2.1) | <2 x 10<sup>-16</sup> |
| Pandemic wave |  |  |
| Before June 30, 2020 | 2.7 (2.7 – 2.8) | <2 x 10<sup>-16</sup> |
| After June 30, 2020 | 1 |  |

<sup>1</sup> Of the entire series, 108,316 cases were excluded due to incomplete or erroneous data, for a total of 4,224,698 SARS-CoV-2-positive subjects analyzed and 109,605 deaths within 30 days of diagnosis. <sup>2</sup> Multivariable Cox analysis (proportional hazards model).

Multivariable analysis of non-hospitalized individuals, 30 days after diagnosis, also showed that age, sex, and pandemic wave were all independent poor prognostic factors **(Table 3)**. Again, age conferred the highest risk of death, with an HR >100 in the groups of subjects 65 years or older and HR >1000 for those 85 years and older. Indeed, the vast majority of deaths (94%) among non-hospitalized patients regarded patients ≥65 years old. The risk estimates associated with male sex and pandemic wave are similar to those for the whole series.

**Table 3.** Factors associated with death in non-hospitalized subjects in the Italian national database ^1^

| Feature | Hazard ratio (95% confidence interval) | P-value <sup>2</sup> |
| --- | --- | --- |
| Age group (years) |  |  |
| 0-14 | 0.3 (0.1 – 0.6) | 5.4 x 10<sup>-4</sup> |
| 15-44 | 1 |  |
| 45-64 | 16.5 (13.6 – 20.1) | <2 x 10<sup>-16</sup> |
| 65-74 | 136.7 (112.6 – 165.9) | <2 x 10<sup>-16</sup> |
| 75-84 | 541.8 (447.3 – 656.3) | <2 x 10<sup>-16</sup> |
| ≥85 | 1537.0 (1269.0 – 1861.0) | <2 x 10<sup>-16</sup> |
| Sex |  |  |
| Female | 1 |  |
| Male | 1.8 (1.8 – 1.9) | <2 x 10<sup>-16</sup> |
| Pandemic wave |  |  |
| Before June 30, 2020 | 3.4 (3.4 – 3.5) | <2 x 10<sup>-16</sup> |
| After June 30, 2020 | 1 |  |

<sup>1</sup> Among non-hospitalized persons, 101,300 cases were excluded due to incomplete or erroneous data, for a total of 3,816,311 SARS-CoV-2-positive subjects analyzed and 27,409 deaths within 30 days of diagnosis. <sup>2</sup> Multivariable Cox analysis (proportional hazards model).

Finally, multivariable Cox analyses of the smaller subgroup of hospitalized patients also showed that age, sex, and pandemic wave were significantly associated with the risk of death 30 days after hospitalization (**Table 4**). The estimates of the risk of death in this subgroup were lower than in non-hospitalized patients, as evidenced by the smaller values of HR not exceeding 100 even in the oldest age group, when compared to the reference group. Finally, being hospitalized during the first wave than later was associated with a higher risk of death (HR= 1.4), but this effect was less intense than that observed among non-hospitalized persons for whom HR = 3.4. Also, the poor prognostic role of male sex was confirmed in these hospitalized patients (HR = 1.5).

**Table 4.** Factors associated with death in hospitalized COVID-19 patients in the Italian national database ^1^

| Feature | Hazard ratio (95% confidence interval) | P-value <sup>2</sup> |
| --- | --- | --- |
| Age group (years) |  |  |
| 0-14 | 0.2 (0.1 – 0.4) | 1.8 x 10<sup>-7</sup> |
| 15-44 | 1 |  |
| 45-64 | 5.5 (5.0 – 6.1) | <2 x 10<sup>-16</sup> |
| 65-74 | 18.2 (16.6 – 20.0) | <2 x 10<sup>-16</sup> |
| 75-84 | 34.5 (31.4 – 37.8) | <2 x 10<sup>-16</sup> |
| 85+ | 58.8 (53.6 – 64.4) | <2 x 10<sup>-16</sup> |
| Sex |  |  |
| Female | 1 |  |
| Male | 1.5 (1.4 – 1.5) | <2 x 10<sup>-16</sup> |
| Pandemic wave |  |  |
| Before June 30, 2020 | 1.4 (1.4 – 1.4) | <2 x 10<sup>-16</sup> |
| After June 30, 2020 | 1 |  |

<sup>1</sup> Among hospitalized persons, 2448 cases were excluded due to incomplete or erroneous data, for a total of 412,942 SARS-CoV-2-positive subjects analyzed and 85,503 deaths within 30 days of diagnosis. <sup>2</sup> Multivariable Cox analysis (proportional hazards model).

## Discussion

The Italian COVID-19 epidemiological surveillance dataset analyzed here contained information on over 4 million persons molecularly diagnosed with a SARS-CoV-2 infection until July 25, 2021. The dataset included information about age, sex, date of diagnosis, presence vs. absence of symptoms, date of hospitalization (if pertinent), date of death (if pertinent), and a few other data. Survival analyses on the whole series and on subsets of non-hospitalized and hospitalized patients strongly confirmed the pivotal role of age in the probability of survival of COVID-19 patients. The analysis by age category, adjusted for sex and pandemic wave, showed that age groups older than 65 had mortality risks that were hundreds of times greater than that of the 15- to 44-year-old reference class. The 0-14 years age group had a mortality risk that was about 10 times less than that of the reference class. Male sex was also confirmed to be a poor prognostic factor, but with a much smaller effect. Additionally, our analysis demonstrated that being diagnosed during the first pandemic wave (until June 2020) was associated with an approximately 3-fold higher mortality risk than being diagnosed later.

In non-hospitalized patients, the mortality risk associated with age was greater than that for the whole series. This difference might be explained by the observation that most deceased non-hospitalized patients were very old, with a median age of 86 years. As a possible interpretation, we suppose that some elderly persons deteriorated rapidly and died before they could be hospitalized.

In hospitalized patients, old age was associated with an excess risk of death, as in the whole series, although the statistical estimates were lower. For example, for the age group ≥85 years old, the HR was 58.8 for hospitalized patients and 692.4 for the whole series. The difference may be explained by the fact that hospitalized patients were much older than subjects of the whole series, i.e., median age 70 years versus 46 years, and that, therefore, hospitalization by itself incorporates an excess risk of death, as age is a known risk factor for hospitalization [15,16], including in our series (not shown).

Our finding of age being a risk factor for COVID-19 mortality is in agreement with that of a meta-analysis by Shi et al. [8] on 27 studies (including 24 from China, two from the United States, and one from Italy) and a meta-analysis by Booth et al. [17] on 66 studies with >17 million patients from 14 countries. Both meta-analyses found an association between old age and excess risk of mortality from COVID-19, although the quantitative risk estimates differ. Of note, these meta-analyses did not report HRs associated with survival, since no Cox analyses were done. To the best of our knowledge, only one other nation-wide study, conducted in France by Semenzato et al. [18], used Cox models to analyze the effects of age on the risk of mortality in a large number of hospitalized COVID-19 patients. Although the age groups differ between the two studies, the risk estimates are similar, with HRs >50 in elderly patients in both studies.

Several immunological mechanisms responsible for the increased risk of death from COVID-19 in the elderly can be hypothesized. One study demonstrated that pre-existing T-cell immunity induced by circulating human alpha- and beta-coronaviruses is present in young adults but virtually absent in older adults [19]. Consequently, older adults had a minimal baseline frequency of cross-reactive T cells directed toward the novel SARS-CoV-2; for this reason, they may be at higher risk of severe COVID-19 disease and death. Moreover, the phenomenon of immunosenescence, which involves age-related changes in innate and adaptive immunity, has been imputed as being associated with the increased mortality of older adults infected with SARS-CoV-2 [20]. The elderly exhibit a deficient immunologic response to SARS-CoV-2 infection, which may be another reason for their increased risk of severe disease and death [21].

In the Italian nationwide COVID-19 series, male sex was an unfavorable prognostic factor for survival, with a risk that was 62% higher than for females in the whole series, and 28% and 27% higher in non-hospitalized and hospitalized patients, respectively. This result is in agreement with those of several other studies [8,17], although the quantitative risk estimates differ. The HRs for male sex calculated in this study, which range from 1.4 to 2.3, are similar to those reported by Semenzato et al. [18].

The mechanism by which sex is an unfavorable prognostic factor for COVID-19 is not yet known. Most likely, several sex-related factors contribute to the higher risk of males for poorer COVID-19 outcomes. A study of 1,683 Italian patients who underwent chest computed tomography at admission showed that men had a higher prevalence of cardiovascular comorbidities, more coronary calcifications, and a higher coronary calcium score than females [22]. Notably, the higher coronary calcific burden of men appeared to be associated with higher mortality. A study of about 3,000 COVID-19 patients in a single center in China observed that the level of inflammatory cytokines in peripheral blood was higher in males than in females [23]. Also, the percentages of CD19+ B cells and CD4+ T cells were generally higher in female patients during the course of the disease. Overall, males had greater inflammation, lower lymphocyte counts, and lower and delayed antibody responses during SARS-CoV-2 infection and recovery than females. Finally, from the perspective of an immunological mechanism, it has been hypothesized that chronic, subclinical, systemic inflammation, characteristic of aging, and immunosenescence contribute to the excess risk of COVID-19 mortality in elderly men [24].

Our multivariable analysis provides strong support for the hypothesis that mortality from COVID-19 was much greater during the first wave (January to June 30, 2020) than later. Indeed, during the first wave, we observed a ∼3-fold excess risk of death in the whole series (HR = 2.7) and in non-hospitalized patients (HR = 3.4) compared to the subsequent period. In hospitalized patient, the excess risk of death was 1.4-fold higher in the first wave than later. The excess risk of death associated with pandemic wave was first reported in an Italian study of hospitalized patients [12], and then confirmed by studies of Massachusetts healthcare workers [13], patients of the U.S. Veterans Affairs healthcare system [25], and UK patients [26]. The reasons for this effect could include the initial lack of preparedness of national health systems for pandemic management, the lack of knowledge about the most effective therapies for COVID-19 patients with severe disease, and the possibility that frailer people were more affected at the beginning of the pandemic than the rest of the population. Another possible explanation of a lower risk of mortality after June 30^th^, 2020, than in the first wave may be the advent of COVID-19 vaccines, from January 2021, that were associated with a reduced risk of mortality [27].

In the Italian COVID-19 epidemiological surveillance dataset, more than 2 million infected persons were symptomatic (50.4% of all cases). Modeling studies on the prevalence of infection in different populations suggested that the total number of SARS-CoV-2-positive individuals exceeds symptomatic cases by an order of magnitude or more [28–30]. If this holds true for the Italian population, then ∼20 million people in Italy have been infected by SARS-CoV-2, i.e., 10 times the 2 million symptomatic cases. Why some infections are asymptomatic and others lead to severe COVID-19 has not yet been elucidated. Cross-reactive immunity, pre-existing in individuals who had been exposed to other coronaviruses, could be one of the mechanisms for asymptomatic and moderate courses of SARS-CoV-2 infection in many individuals [31].

A limitation of our study is the lack of data about COVID-19 patients’ comorbidities, which are important risk factors for outcome [8]. This lack of information prevented us from analyzing other risk factors for death. Moreover, the reasons why some hospitalized patients were classified as asymptomatic are not known, but their hospitalization may have been due to reasons other than COVID-19. For example, in 5,432 cases, the date of SARS-CoV-2 infection detection was after the date of hospitalization and in 13,144 patients the diagnosis was on the same day.

Overall, this study confirms that age and male sex are independent risk factors for COVID-19 mortality for both hospitalized and not-hospitalized patients. Because age was found to be the most impactful negative prognostic factor, it should be considered in pandemic management, by giving priority to strategies aimed at protecting elderly people. Additionally, this is the first country-wide study to demonstrate a high risk of mortality during the first pandemic wave than later. Similar nation-wide studies in different countries, to the best of our knowledge, have not been published. Thus, we cannot compare our study with those from other nations with different mortality rates, and we cannot exclude that such differences are due to unequal pandemic management in the first wave, considering that Italy was the first Western nation to be affected. Our study also suggests that the medical research that started with the pandemic onset and that led to the development of increasingly more effective clinical protocols contributed to improving COVID-19 patient survival. Despite the limitations of this study, principally due to the lack of some clinical data (e.g. about comorbidities), this study demonstrates the usefulness of a national database for studying a new disease such as COVID-19. Efforts should be made in Italy to create a more detailed national database like those of the United Kingdom [32] and France [33] that collect more data on demographics, symptoms, diagnostic tests and treatments. National health databases, especially when accompanied by a national biobank of blood samples, offer great possibilities for biomedical research. They allow the construction of cohorts with unparalleled statistical power and help study risk factors for common diseases, rare diseases, and new emerging diseases such as COVID-19. Their availability could impact treatment and public health. Therefore, the creation of such databases in countries that do not yet have them and the creation of European databases are desirable.

## Data Availability

ISS data that have been used for the present study are available upon request at this web link: https://www.iss.it/richiesta-dati-covid19

## Ethical statement

Data used in this study were collected by the Istituto Superiore di Sanità (Rome, Italy) for the Italian national integrated COVID-19 surveillance. The scientific dissemination of these data was authorized by the Italian Presidency of the Council of Ministers on February 27, 2020 (Ordinance n. 640) and August 4, 2020 (Ordinance n. 691). Data were provided to us upon request (protocol no. AOO-ISS-19/04/2921-0014810).

## Acknowledgments

The authors acknowledge the contributions of Valerie Matarese, PhD, who provided scientific editing.

